# Mindfulness-based intervention for very preterm young adolescents: An RCT

**DOI:** 10.1101/2021.03.14.21253449

**Authors:** Vanessa Siffredi, Maria Chiara Liverani, Petra Susan Hüppi, Lorena Freitas, Jiske De Albuquerque, Fanny Gimbert, Arnaud Merglen, Djalel-Eddine Meskaldji, Cristina Borradori Tolsa, Russia Hà-Vinh Leuchter

## Abstract

**Objectives:** This randomised controlled trial (RCT) assesses the effectiveness of a Mindfulness-Based Intervention (MBI) in improving executive, behavioural and socio-emotional competences in very preterm young adolescents.

**Methods:** 58 young adolescents aged 10 to 14 years, born before 32 gestational weeks at the Geneva University Hospital, Switzerland, participated in the study. They were randomly assigned to an “intervention” or a “waiting” group and all completed an 8-week MBI in a cross-over design. Executive, behavioural and socio-emotional competences were assessed at three different time points via parent and self-reported questionnaires, neuropsychological testing, and computerised tasks. We analysed data using an intention-to-treat approach with linear modelling. Subgroups of participants based on levels of prematurity were created using k-means clustering.

**Results:** Parent questionnaires revealed a statistically significant immediate effect of MBI with increased executive and behavioural competencies in everyday life. Increased executive competence was also observed on a Flanker task with enhanced speed of processing after MBI. Two subgroups of participants were created based on measures of prematurity, which revealed increased long-term benefits in the moderate-risk that are not observed in the high-risk subgroups of VPT young adolescents.

**Conclusions:** Our findings show a beneficial effect of MBI on executive, behavioural and socio-emotional competences in VPT young adolescents. Moderate-risk and high-risk VPT young adolescents showed different immediate and long-term beneficial effects of the intervention. Our results suggest that MBI is a valuable tool for reducing detrimental consequences of prematurity in young adolescents, especially regarding executive, behavioural and socio-emotional difficulties.

**Data Sharing Statement:** Deidentified individual participant data (including data dictionaries) will be made available, in addition to study protocols, the statistical analysis plan, and the informed consent form. The data will be made available upon publication to researchers who provide a methodologically sound proposal for use in achieving the goals of the approved proposal. Proposals should be submitted to.

## INTRODUCTION

Follow-up studies indicate that VPT individuals are at increased risk for executive, behavioural and socio-emotional difficulties in childhood that persists into adolescence and adulthood. ^1-13^ According to the model of Anderson (2002), executive functioning (EF) is essential for goal-directed and adaptive problem-solving and behaviour and it is conceptualised in four distinct subdomains: (i) attentional control, (ii) information processing, (iii) cognitive flexibility, and (iv) goal setting ^14^. On the other hand, behavioural and socio-emotional competences refer to a set of skills related to how individuals identify, express, understand, use and regulate their behaviour as well as their emotions and those of others. ^15^ Importantly, these competences are crucial in daily life activities, with a close link to academic abilities and significant implications on social behaviour.^16-19^

These findings suggest that VPT children and adolescents may benefit from interventions designed to enhance executive, behavioural and socio-emotional competences. In recent years, general interest in the practice and benefits of mindfulness-based interventions (MBI) has increased. Mindfulness is commonly defined as the on-going monitoring of present-moment experience while attending to it with openness, nonjudgment and acceptance.^20^ Despite underlying mechanisms are still to be further explored, numerous studies conducted in children and adolescents have shown that MBI can be a valid way to support the development of executive functions, including attentional control and information processing speed, as well as behavioural and socio-emotional competences, such as emotion regulation. ^21-29^

This randomised controlled trial (RCT) aims to assess the effectiveness of an 8-week MBI in VPT young adolescents aged 10 to 14 years to improve executive, behavioural and socio-emotional functioning. The age of 10 to 14 years has been targeted as a crucial developmental period during which MBI may be beneficial.^30^

## METHODS

The “Mindful preterm teens” study is an RCT of an MBI in VPT adolescents aged 10 to 14 years (Swiss Ethics Committees on research involving humans, ID: 2015-00175), see Siffredi, Liverani and colleagues for a detailed description.^31^ Written informed consent was obtained from primary caregivers and participants.

### Participants

One hundred and sixty-five VPT young adolescents were invited to participate in the study. They were aged 10 to 14 years, born before 32 gestational weeks between 01.01.2003 and 31.12.2008 in the Neonatal Unit at the Geneva University Hospital, Switzerland, and received follow-up care at the Division of Child Development and Growth at the Geneva University Hospital. VPT young adolescents were excluded if they had an intelligence quotient below 70, sensory or physical disabilities (cerebral palsy, blindness, hearing loss), or an insufficient understanding of French. Moreover, some families declined to participate due to lack of time, lack of interest, geographical constraints or unreachability. Out of the 165 young adolescents invited to participate, 56 (33.9%) were enrolled in the RCT, see Figure 1.

**Figure 1.**
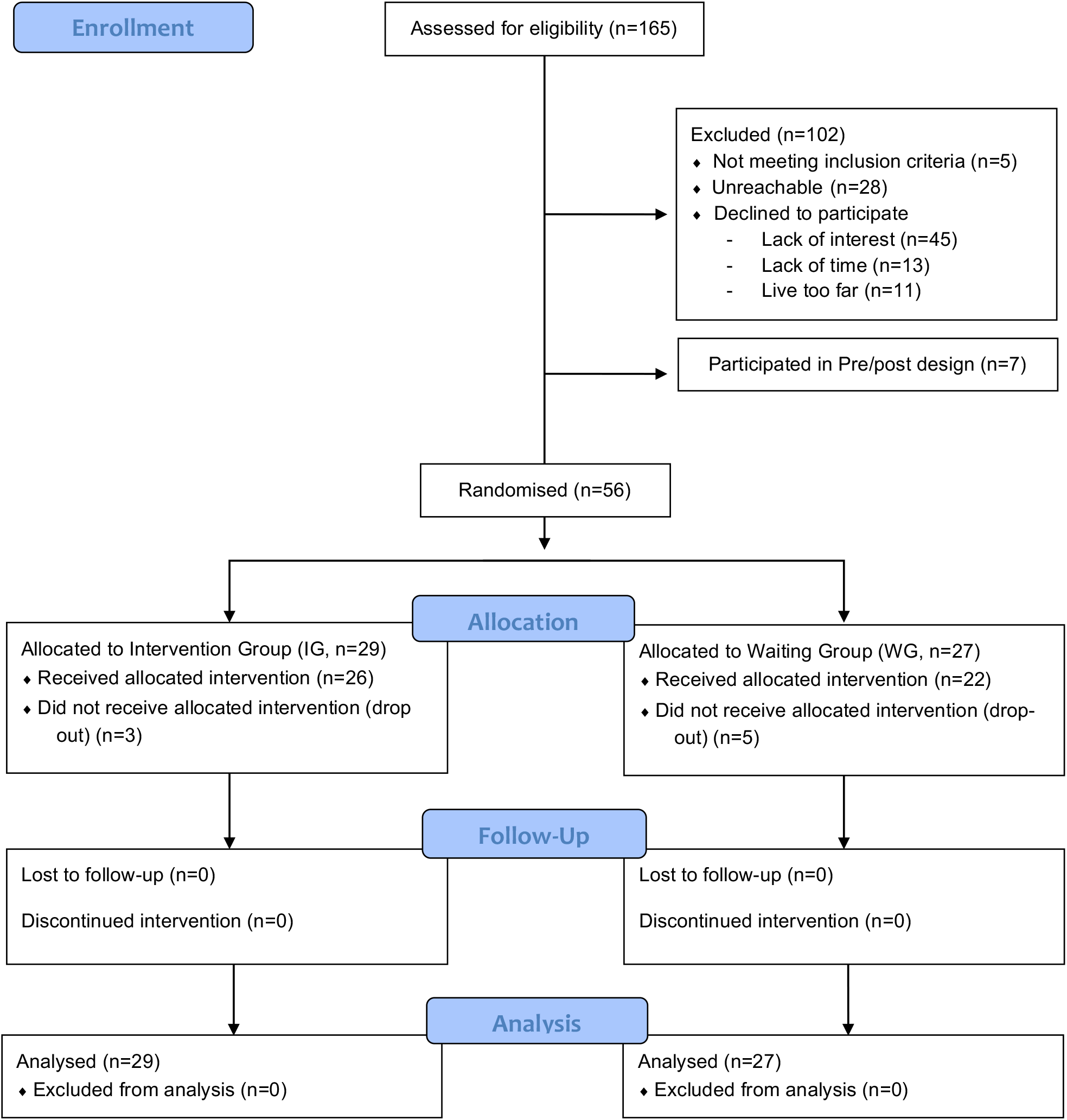
CONSORT flow diagram of the present cross-over RCT design

### Procedures

Once enrolled in the RCT, families were allocated to the intervention group (IG) or the waiting group (WG) with a cross-over RCT design, see Figure 2. All participants completed a baseline assessment to evaluate general intellectual functioning and demographic characteristics. Additional assessments were completed at three different time points, where outcome measures were collected via parent-report and self-report questionnaires, neuropsychological assessments and computerised neurocognitive tasks. Children from the IG completed the MBI between Time 1 and Time 2. Participants from the WG completed the MBI between Time 2 and Time 3. For all young adolescents involved in the trial, the pre-intervention assessment (i.e., Time 1 for the IG, and Time 2 for the WG) was completed within one month before the first MBI session. The post-intervention assessment (i.e., Time 2 for the IG, and Time 3 for the WG) was completed within one month after the last MBI session. For the IG, the remaining assessment (i.e., Time 3) was completed three months after the post-intervention assessment and will be referred to as “Long term” assessment. For the WG, the remaining assessment (i.e., Time 1) was completed three months before the pre-intervention assessment.

**Figure 2.**
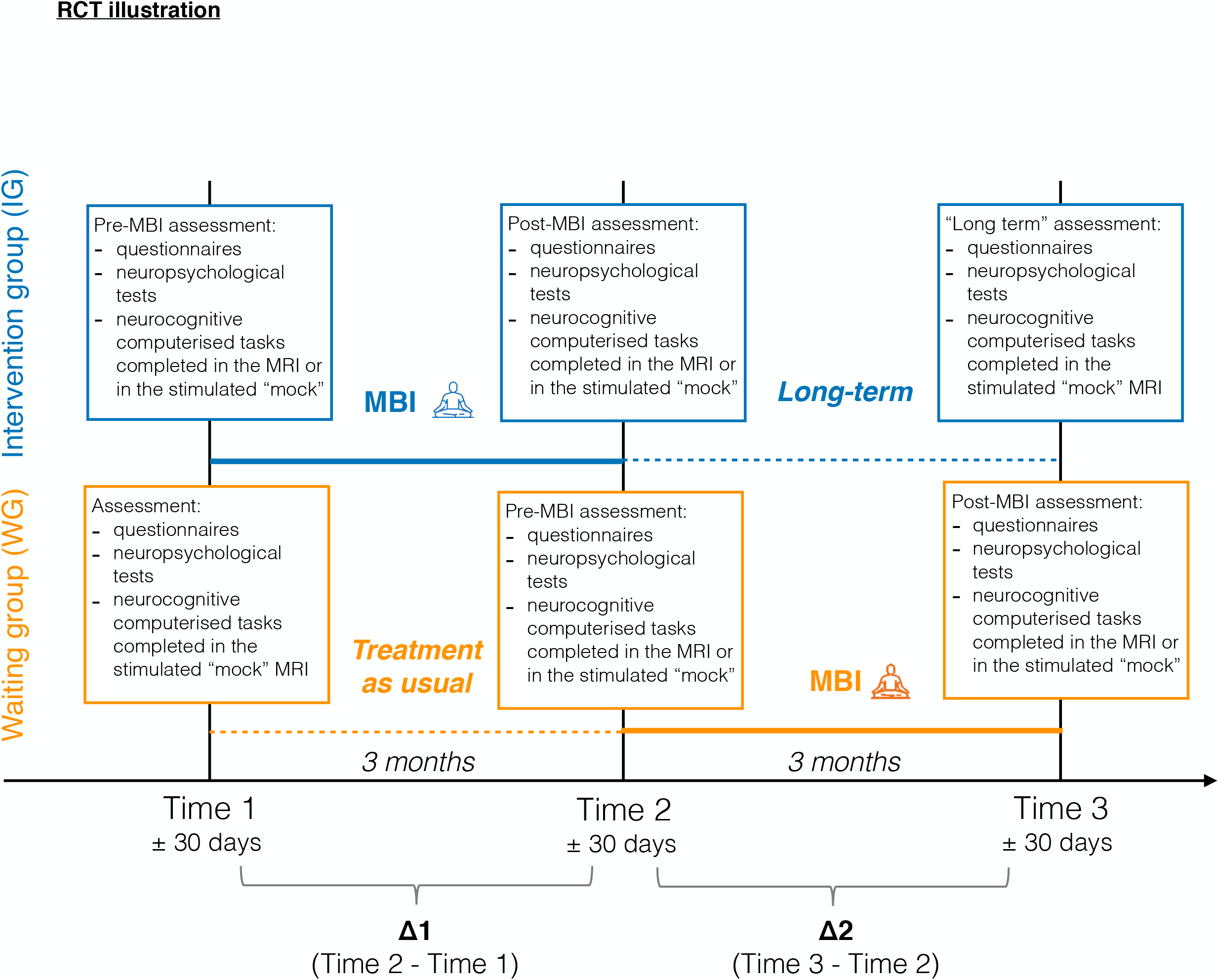
Illustration of the RCT study design. Participants enrolled in the RCT design were randomised in two groups: the intervention group (IG) in blue and the waiting group (WG) in orange.

### Mindfulness-based Intervention

MBI consisted of eight weekly sessions in groups of up to seven participants, lasting ninety minutes, as well as an invitation to practice daily at home. Two instructors were present for each group throughout the intervention. The MBI program used in this study was specifically adapted to adolescents, see Supplementary Methods.

### Neonatal and Demographic Characteristics

Neonatal characteristics were documented from medical records. In order to estimate general intellectual functioning, the General Ability Index (GAI) from the Wechsler Intelligence Scale for Children – 4th Edition (WISC-IV)^32^ was used. Parent-report and self-report demographic questionnaires were used to assess general characteristics of the participant. Socio-economic status was estimated from maternal education and paternal occupation using the validated Largo scale. Higher socio-economic scores reflect lower socio-economic status levels.^33^

### Outcome measures

Participants’ executive, behavioural and socio-emotional functioning were assessed using parent-report and self-report questionnaires, neuropsychological testing and computerised neurocognitive tasks, see supplementary Table S1.

#### (i) Executive competences measures

Executive competences of young adolescents were assessed using the Behaviour Rating Inventory of Executive Function – parent version (BRIEF),^34^ evaluating attention, hyperactivity and impulsivity in everyday life. The BRIEF comprises 86 items over two standardised subscales, the Behavioural Regulation Index (BRI) and the Metacognition Index (MI), as well as a global score called the Global Executive Composite (GEC). Neurocognitive computerised tasks comprised: (i) the Flanker Visual Filtering Task, in which reaction time of the congruent condition was used to assess speed of processing, which belongs to the information processing subdomain, and the inhibition score (reaction time in incongruent conditions – reaction time in congruent conditions) was used as a measure of the attentional control subdomain;^14, 35^ (ii) the child-adapted version of the Reality Filtering task, in which the temporal context confusion index (TCC) was used as a reality filtering measure, which involves integration of different executive processes.^36, 37^ Neuropsychological testing included the Letter-Number Sequencing subtest from WISC-IV assessing working memory, which belongs to the cognitive flexibility subdomain.^14^ Given the strong association between executive functions and mathematical abilities in children and adolescents,^38, 39^ we also used the total score of the Tempo Test Rekenen to assess timed mathematical achievement.^40^

#### (ii) Behavioural and socio-emotional competences measures

The total score of the Strength and Difficulties Questionnaire – parent version (SDQ) was used to assess behaviour in daily life.^41, 42^ Participants completed three self-reported questionnaires: the KIDSCREEN-27 items questionnaire was used to assess the quality of life of the participants;^43^ the total score of the Social Goal Scale was used to assess social responsiveness and social relationships;Wentzel ^44^ and the total score of the Self-Compassion Scale – Short form was used to assess the main components of self-compassion.^45^ Neuropsychological testing included the Affect Recognition subtest (NEPSY-II), giving a total score assessing facial emotional recognition,^46^ and the Theory of Mind subtest (NEPSY-II), giving a total score measuring the ability to understand mental functions, such as belief, intention or deception.

### Statistical Analyses

#### Main statistical analyses

All analyses were based on the intention-to-treat principle. For each outcome measure, raw scores were used to calculate differences between Time 1 and Time 2 (Time 2-Time 1 = Δ1), and between Time 2 and Time 3 (Time 3 – Time 2 = Δ2) for each participant, see Figure 2. Negative Δ indicates a reduction of the scores between two time points, whereas positive Δ indicates an increase in scores between two time points. Linear models were used to evaluate the effect of MBI. Assumptions of linear models were assessed based on visual diagnosis of the distribution of the residuals. We modelled fixed effects of outcome measures as dependent variables and interaction of time (i.e., Δ1 and Δ2) by group (i.e., IG and WG) as independent variables. When the model’s p-value was significant, we used planned contrasts to compare outcome measures between the different levels of the independent variables time and group:

- we assessed the effect of the intervention immediately after MBI using the planned contrast defined as: “MBI” (i.e., Δ1 of IG and Δ2 of WG) versus “treatment as usual” (i.e., Δ1 of WG).
- we assessed delayed effect of MBI using the planned contrast defined as: “long-term” (i.e., Δ2 of IG) versus “treatment as usual” (i.e., Δ1 of WG).
- when the effect of the intervention immediately after MBI was significant (“MBI” versus “treatment as usual”), we assessed the long-term effect of the intervention using the planned contrast defined as: “MBI” (i.e., Δ1 of IG and Δ2 of WG) vs “long-term” (i.e., Δ2 of IG).

Effect size and p-values were calculated. The p-values were also corrected for multiple comparisons using the Benjamini and Hochberg method (1995), which controls the False Discovery Rate correction (FDR, q-values ≤ 0.05).^47^ All analyses were performed using R software, version 3.5.2.^48, 49^

#### Subgrouping “Prematurity” analyses

In order to better understand inter-individual differences, we performed exploratory analyses on specific subgroups of VPT pre-adolescents. Clustering analyses were used to explore whether any treatment effect tested in our RCT varied across subgroups defined by pre-intervention patient characteristics.^50^ Subgrouping of participants was determined by K-means clustering and was based on the main properties of premature birth. A subgrouping “prematurity” was created by using the measures of birth weight and gestational age as features to create two groups of VPT participants: the “high-risk” group, including participants with lower birth weight and lower gestational age, and the “moderate-risk” group, including participants with higher birth weight and higher gestational age. To evaluate the effect of MBI on these subgroups, analyses similar to the section above were conducted.

## RESULTS

### Neonatal and demographic characteristics

Neonatal and demographic characteristics of the 56 participants enrolled in the RCT are shown in Table 1. There were no significant differences in demographic and clinical characteristics at the age of 10-14 years between IG and WG (gender, age, index of general cognitive ability and socio-economic status) and the neonatal characteristics between IG and WG (gestational age, head circumference, length of hospitalisation, presence of severe brain lesions and other medical conditions).

**Table 1.** Neonatal and demographic characteristics at baseline of young adolescents enrolled in the RCT (n=56), as well intervention group (IG) and waiting group (WG) comparisons

|  |  | RCT, n=56 |  |  |
| --- | --- | --- | --- | --- |
|  |  | Intervention group (IG),<br>n=29 | Waiting group (WG),<br>n=27 | Group comparison<br>(IG vs WG) |
| **Neonatal characteristics** |  |  |  |  |
| Birth weight, mean (SD) [range] in grams |  | 1284.83 (351.41) [650;1810] | 1210 (400.85) [520;1980] | t(54)=0.744, p=0.460 |
| Gestational age, mean (SD) [range] in days |  | 29.29 (1.92) [24.71; 31.86] | 29.12 (1.93) [26;31.71] | t(54)=0.317 p=0.753 |
| Head circumference, mean (SD) [range] in cm |  | 26.55 (2.57) [21;31] | 25.65 (2.82) [21;31] | t(53)=1.234, p=0.223 |
| Length of hospitalisation, mean (SD) [range] in days |  | 59.56 (26.79) [23;131] | 63 (33.69) [17;151] | t(52)=-0.416,p=0.679 |
| Multiple births, n (%) | | 13 (44.8%) | 7 (25.9%) | $\chi^2(2)=2.202$, p=0.333 |
| cPVL, n(%) | | 1 (3.4%) | 0 | $\chi^2(1)=0.903$, p=0.342 |
| IVH - Grades III and IV, n (%) |  | 0 (0%) | 0 (0%) | - |
| BPD, n (%) | | 5 (17.2%) | 6 (22.2%) | $\chi^2(1)=0.534$, p=0.465 |
| **Demographic characteristics** |  |  |  |  |
| Gender | Female, n | 14 (48.3%) | 16 (59.3%) | $\chi^2(1)=0.678$, p=0.410 |
|  | Male, n | 15 (51.7%) | 11 (40.7%) |  |
| Age at baseline, mean (SD) [range] in years |  | 12.05 (1.23) [10.08;14.24] | 12.26 (1.37) [10.38;14.85] | t(50)=-0.585, p=0.561 |
| Index of general ability (GAI), mean (SD) [range] |  | 106.67 (11.47) [83;132] | 108.76 (11.23) [87;130] | t(50)=-0.664, p=.0.510 |
| Socio-economic status (SES), mean (SD) [range] |  | 4.78 (2.62) [2;12] | 3.76 (2.35) [2;12] | t(50)=1.470, p=0.148 |

Abbreviations: Cystic Periventricular Leukomalacia = cPVL, Intraventricular haemorrhage = IVH, Bronchopulmonary dysplasia = BPD. Note: Independent-sample t-test, Chi-square was used to compare the randomised groups

### RCT timing

Time differences (in days) between Time 1 and Time 2, as well as between Time 2 and Time 3 were not significantly different between the IG and the WG (p=0.496, p=0.502), see supplementary Table S2.

### Main Outcomes

#### Executive competences outcomes

Planned contrasts “MBI” vs “treatment as usual” showed a significant effect of the MBI on the BRIEF GEC and MI delta scores, reflecting enhanced executive capacities in everyday life (p=0.002 and p<0.001 respectively). This beneficial effect on executive functioning was supported by a significant decrease in delta reaction time on the processing speed measure of Flanker task (p<0.001). Planned contrasts “MBI” vs “long-term” showed a significant increase for both BRIEF GEC and MI delta scores (p=0.008 and p=0.002), showing that the beneficial effect of MBI was not maintained three months after the end of the intervention. The planned contrast “treatment as usual” vs “long-term” showed a significant decrease in reaction time on the Flanker task processing speed measure (p=0.01), reflecting a long-lasting effect of the MBI on this information processing subdomain, Figure 3. There was no robust effect on other executive scores, Supplementary Tables S3.

**Figure 3.**
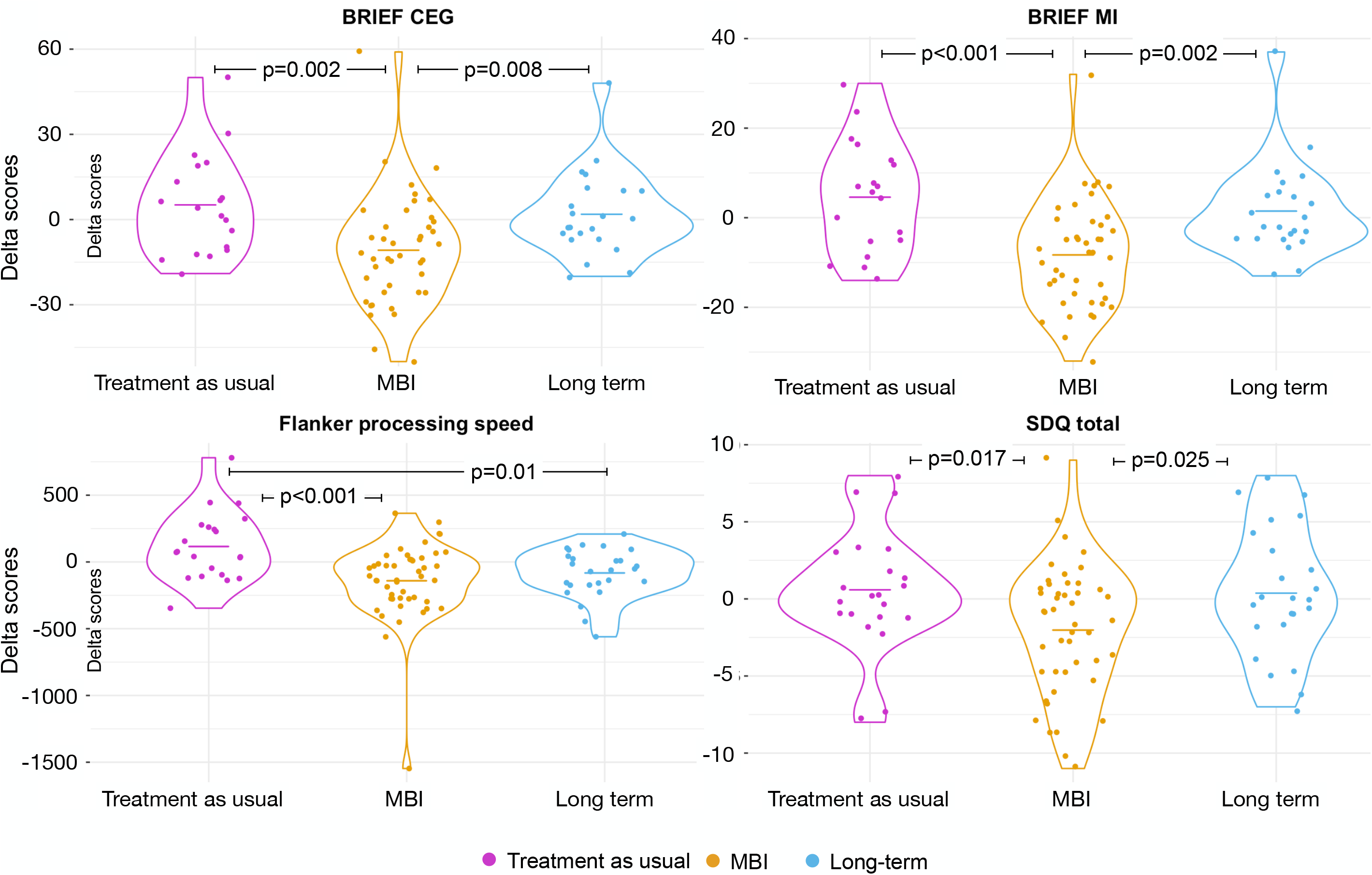
Plots showing the distribution of the delta scores (Δ) of the “Treatment as usual”, “MBI” and “Long-term” groups of the significant planned contrasts only. Lines in the violin plots represent the means for each group.

#### Behavioural and socio-emotional competences measures

The planned contrast “treatment as usual” vs “MBI” showed a significant effect of the MBI on the SDQ delta total score with a significant decrease in scores after MBI (p=0.017), reflecting an improvement in general behavioural competences, Figure 3. The planned contrast “MBI” vs “long-term” showed a significant increase in SDQ delta total score, showing that the beneficial effect of MBI was not maintained three months after the end of the intervention. There was no robust effect for the quality of life and socio-emotional competencies, Supplementary Tables S4.

#### Subgrouping “Prematurity”

Using K-means clustering, two groups of VPT participants were extracted based on weight and gestational age at birth: the high-risk group [n=29, gestational age: mean (SD)= 27.91 (1.62); birth weight: mean (SD)= 938.1 (197.08)] and the moderate-risk group [n=27, gestational age: mean (SD)= 30.63 (0.91); birth weight: mean (SD)= 1583.89 (196.8)].

#### Executive competences outcomes

Planned contrasts “treatment as usual” vs “MBI” showed a significant effect of the MBI in both the high- and moderate-risk subgroups for the BRIEF MI (high-risk, p=0.016; moderate-risk, p=0.003) with a significant decrease of BRIEF MI delta scores; as well as a decrease in BRIEF GEC deltas scores only in the high-risk subgroup (p=0.011). The planned contrasts “MBI” vs “long-term” and “treatment as usual” vs “long-term” showed a significant increase in the BRIEF MI and CEG delta scores three months after MBI in the high-risk subgroup only, reflecting that the beneficial effect of MBI was not maintained in this group, Figure 3. For both subgroups, planned contrasts “treatment as usual” vs “MBI” showed a significant decrease in delta reaction time on the Flanker task, reflecting increased processing speed after MBI (high-risk, p=0.035; moderate-risk, p=0.001). In the moderate-risk subgroup only, planned contrasts “treatment as usual” vs “long-term” showed a significant decrease in reaction time on the Flanker task, reflecting an increase in processing speed that lasted three months after the end of the MBI (p=0.001), Figure 3. There was no robust effect for the other executive scores, Supplementary Tables S5 and S6.

#### Behavioural and socio-emotional competences outcomes

For significant linear models adjusted for multiple comparisons, planned contrasts “treatment as usual” vs “MBI” showed a significant increase in self-compassion delta scores after MBI specific to the high-risk subgroup (p=0.004), reflecting enhanced self-compassion after MBI, Figure 4. For both the moderate- and the high-risk subgroups, planned contrasts “treatment as usual” vs “long-term” showed a significant increase in self-compassion scores three months after the end of the intervention (moderate-risk, p=0.002; high-risk, p=0.008). There was no robust effect for the behavioural and quality of life scores, Supplementary Tables S5 and S6.

**Figure 4.**
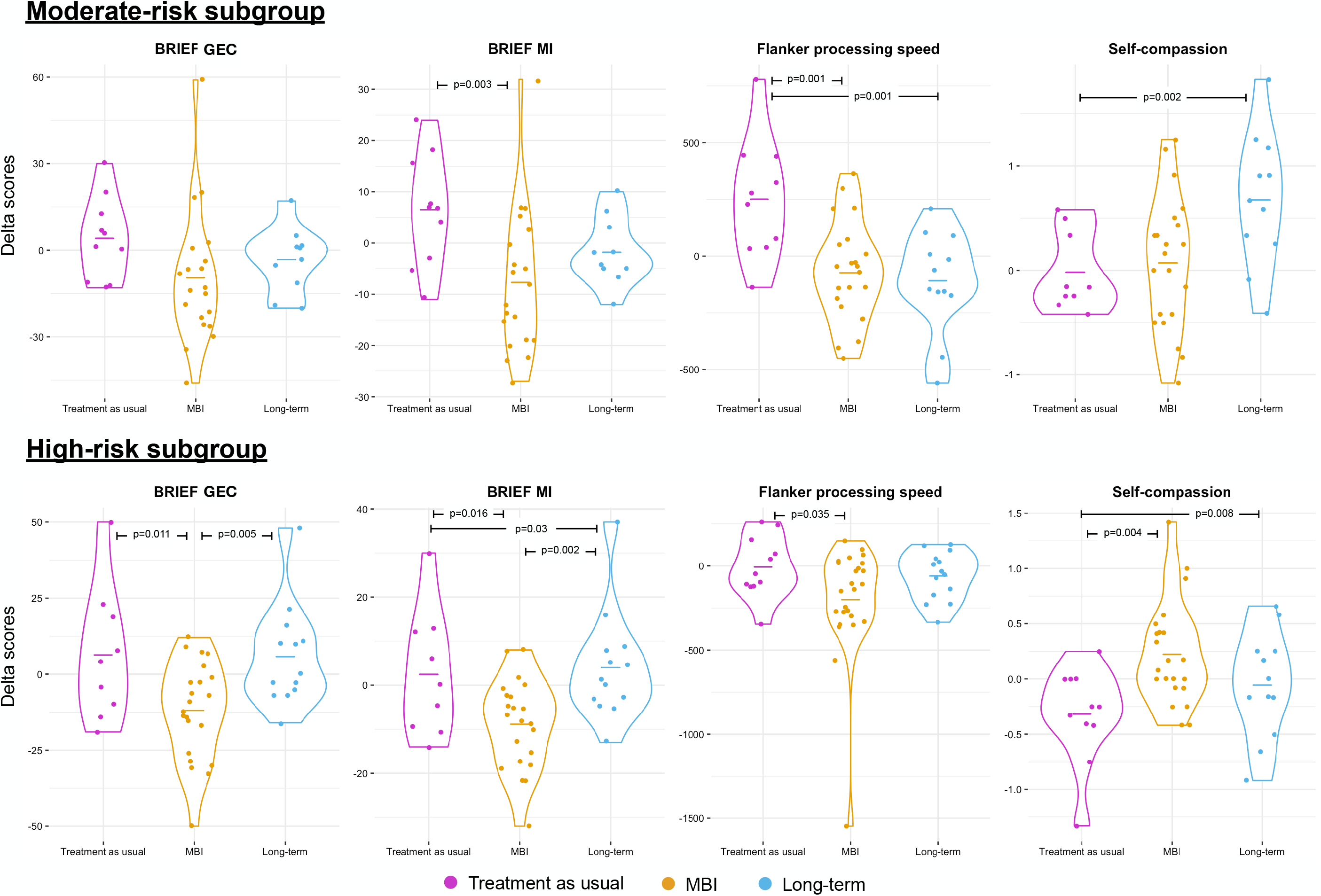
Distribution of the delta scores (Δ) of the “Treatment as usual”, “MBI” and “Long-term” groups for the significant post-hoc tests for the two subgroups of VPT: moderate-risk and high-risk. Lines in the violin plots represent the means for each group.

## DISCUSSION

This RCT assessed the effectiveness of an 8-week MBI in VPT young adolescents to improve executive, behavioural and socio-emotional competences. Our findings show beneficial effects of MBI immediately after the intervention on executive, behavioural and behavioural competences in every-day life based on parent-reported questionnaires and on processing speed capacities. Subgrouping analyses based on the level of prematurity reveal a larger beneficial effect of MBI immediately after the intervention in the high-risk VPT subgroup, but larger long-lasting effects of the MBI in the moderate-risk VPT subgroup. Our findings lead us to conclude that the use of MBI in VPT young adolescents is effective in improving behavioural as well as executive and socio-emotional outcomes.

Parent-report questionnaires revealed an increase in overall executive competences in everyday life, together with a more specific effect on metacognitive abilities. An enhancement of processing speed on a computerised task corroborates these results reflecting increased skills in the information processing EF subdomains.^14^ These findings are in line with previous studies conducted in different populations of children and adolescents showing strong effect of MBI on processing speed.^51-54^ Although we found a long-lasting beneficial effect of MBI three months post-intervention on processing speed capacities, the beneficial effect of MBI on overall executive and behavioural competences reported by parents, was not maintained. Subgrouping analyses based on prematurity levels gave valuable insight into these results. In fact, regarding executive competences, the high-risk subgroup appears to benefit slightly better from the MBI immediately post-intervention, with greater enhancement of overall executive competences in daily life, in addition to improvements in metacognitive abilities and processing speed compared to the moderate-risk group. Nevertheless, the decline in executive competences observed three months post-MBI seems mostly driven by the high-risk subgroup. At the opposite, the long-lasting effect of MBI on processing speed was found only in the moderate-risk group.

When exploring behavioural and socio-emotional competences, our results showed a significant improvement immediately after MBI only on the total score of the SDQ parent-reported questionnaire, reflecting an improvement in general behaviour, but not in the other questionnaires and neuropsychological testing assessing socio-emotional competences. These findings are in line with previous research showing enhancement of behavioural competences after MBI during adolescence.^55-57^ Nevertheless, this effect was not maintained three months after the end of the intervention. In regards to self-compassion (self-reported questionnaire), the subgrouping analyses revealed a significant improvement immediately after the MBI only in the high-risk VPT group. In contrast, a significant improvement three months after the end of the MBI was observed in both the high- and moderate-risk groups.

Our study has several strengths. We used gold standard RCT design, recruited a relatively large sample of VPT young adolescents and analysed the data on an intention-to-treat basis. Nevertheless, theoretical and methodological limitations of this study should inform future research. First, the beneficial effect of MBI observed via parent-reported questionnaires might be questionable given the subjective aspect of these tools.^58^ Future studies should consider the completion of questionnaires by multiple informants from different settings (e.g. parents and teachers) to give a more objective view of the changes occurring after MBI.^55, 59^ Second, one of the main study limitations is the absence of an active control condition or a placebo condition. This would allow participants and their families to be blinded to treatment allocation, as well as help understand what effects are specifically attributable to MBI. Finally, factors such as home environment, caregiver involvement, and motivation to participate in the training and quantity of home practice were not considered in our study.^60, 61^This might influence the outcomes of an MBI and should be considered in future research.

## CONCLUSION

In conclusion, this study shows for the first-time beneficial effects of MBI in VPT young adolescents on enhancing executive, behavioural and socio-emotional competences. Subgrouping analyses based on prematurity level reveal a larger beneficial effect of MBI immediately post-intervention in the high-risk subgroup, but a larger long-lasting effect of MBI in the moderate-risk subgroup. We conclude that the use of MBI in VPT young adolescents is effective in improving executive, behavioural and socio-emotional outcomes. However, a longer MBI intervention might be beneficial for high-risk VPT young adolescents. Although future investigations are needed, MBI seems a promising tool to enhance executive, behavioural and socio-emotional outcomes in a vulnerable population such as VPT young adolescents.

## Data Availability

Deidentified individual participant data (including data dictionaries) will be made available, in addition to study protocols, the statistical analysis plan, and the informed consent form. The data will be made available upon publication to researchers who provide a methodologically sound proposal for use in achieving the goals of the approved proposal. Proposals should be submitted to.

## ACKNOWLEDGMENTS

We thank and acknowledge all participating young adolescents and families who made this research possible. We also thank the Fondation Campus Biotech Geneva (FCBG), a foundation of the Swiss Federal Institute of Technology Lausanne (EPFL), the University of Geneva (UniGe), and the University Hospitals of Geneva (HUG); the Research Platform of the University Hospitals of Geneva (HUG) for their practical help; as well as Mariana Magnus Smith and Françoise Stuckelberger-Grobéty for their implication as MBI instructors.

## Contributors’ Statement

Dr Siffredi et Dr Liverani collected data, coordinated and supervised data collection, carried out the statistical analyses, drafted the initial manuscript, reviewed and revised the manuscript.

Professor Hüppi conceptualized and designed the study, reviewed and revised the manuscript, provided funding.

Dr Freitas collected data, reviewed and revised the manuscript.

Dr De Albuquerque collected data, coordinated and supervised data collection, reviewed and revised the manuscript.

Dr Gimbert collected data, coordinated and supervised data collection, reviewed and revised the manuscript.

Dr Merglen conceptualised and designed the study, reviewed and revised the manuscript. He was one of the instructors of the MBI intervention

Dr Borradori Tolsa conceptualised and designed the study, coordinated and supervised data collection, reviewed and revised the manuscript.

Dr Meskaldji supervised statistical analyses, reviewed and revised the manuscript.

Dr Hà-Vinh Leuchter conceptualised and designed the study, coordinated and supervised data collection, supervised statistical analyses, reviewed and revised the manuscript. She was one of the instructors of the MBI intervention.

All authors approved the final manuscript as submitted and agree to be accountable for all aspects of the work.

## Abbreviations

RCT: Randomised Controlled Trial
MBI: Mindfulness-Based intervention
VPT: Very Preterm
IG: Intervention Group
WG: Waiting Group
WISC-IV: Wechsler Intelligence Scale for Children
GAI: General Ability Index
BRIEF: Behaviour Rating Inventory of Executive Function, parent version
BRI: Behavioural Regulation Index from the BRIEF
MI: Metacognition Index from the BRIEF
GEC: Global Executive Composite from the BRIEF
TCC: Temporal Context Confusion index from the Reality Filtering task
SDQ: Strength and Difficulties Questionnaire, parent version

