## Supplementary Materials for "Mindfulness-based intervention for very preterm young adolescents: An RCT"

**Supplementary Methods**

### Mindfulness-based Intervention

The MBI program used in this study was specifically adapted to adolescents. For each session one theme was addressed, such as attention and the stabilisation of the focus of attention, bodily sensations, breath, emotions, thoughts, compassion, stress, stress reactivity and coping strategies. Different formal meditation practices were introduced, such as sitting mediation with different objects of attention, body scan, walking meditation and mindful movement. Participants had the opportunity to share experiences with their peers and the instructors during each session, allowing the recognition of individual patterns of behaviour, and at the same time, the appreciation of the shared experiences as a community. They were also invited to practice between 5 to 20 minutes per day at home, using guided mediation recorded by the instructors, see Siffredi*, Liverani* & al., under revision, provided in supplementary file.

**Supplementary Table S1**. Details of the RCT outcome measures

| Domains | Modalities | Measures | Description | Scores |
|---|---|---|---|---|
| **Executive competences** | | | | |
| | Parent questionnaire | | | |
| | | Behaviour Rating Inventory of Executive Function, parent version (BRIEF, Gioia, Isquith, Guy, and Kenworthy (2000)) | | |
| | | | The BRIEF parent questionnaire provides an index of attention, hyperactivity and impulsivity in everyday life. The BRIEF comprises 86 items over two standardised subscales: (i) Behavioural Regulation Index (BRI) comprising 3 subscores including, Inhibit, Shift, Emotional Control; (ii) Metacognition index (MCI) comprising 5 subscores including, Initiate, Working Memory, Plan/Organise, Organisation of Materials, | BRIEF CEG<br>BRIEF BRI<br>BRIEF MCI |

| | | |
|---|---|---|
| | Monitor; as well as a global score called the Global Executive Composite (GEC). | |
| | Neuropsychological tests | |
| | Letter-Number Sequencing (WISC-IV, Wechsler (2003)) | |
| | The letter-number sequencing is a working memory task. Sequences of number and letters are read to the participant, and he/she is then asked to re-sequence the numbers in numerical order from lowest to highest and then to sequence the letters in alphabetical order. Standardised scores were used. | Letter-number sequencing |
| | Tempo Test Rekenen (De Vos, 1992) | |
| | The Tempo Test Rekenen is an arithmetic test consisting of 200 arithmetic number fact problems presented in five rows (one row with addition, one row with subtraction, one row with division, one row with multiplication, and one mixed problem row). Within each row, the problems increase in difficulty. Participant are asked to solve as many items as possible within 1 min per row. The total raw score was age-adjusted for each participant using the procedure described in the main statistical analyses section. | Tempo test |
| | Neurocognitive computerised tasks | |
| | Flanker Visual Filtering Task (Christ, Kester, Bodner, & Miles, 2011) | |
| | The Flanker Visual Filtering Task was used to assess attentional control and information processing speed. Each trial showed a horizontal row of five fish. The participant was asked to respond as quickly as possible to whether the central fish was facing to the left or right. Congruent trials were the ones with all five fish in the horizontal row pointing in the same direction and incongruent trials were the ones with the four distracting fishes pointing in the opposite direction of the central target fish. Reaction time of the congruent condition and of the incongruent condition were used to assess information processing speed, and the inhibition score (reaction time in incongruent conditions – reaction time in congruent conditions) was used as a measure of attentional control. | -Flanker processing speed<br>-Flanker inhibition |
| | Reality Filtering Task (Liverani et al., 2017; Schnider, 2018) | |
| | The Reality Filtering task child-adapted version was used to assess recognition memory and orbitofrontal reality filtering. It consisted of a continuous recognition task composed of two runs with the same picture set but arranged in different order. Accuracy of the second run (D2) and Temporal Context Confusion index (TCC as defined by Schnider, 2018) measures reality filtering. | Reality filtering TCC |
| | **Behaviour and socio-emotional competences** | |
| | Parent questionnaire | |
| | Strength and Difficulties Questionnaire, parent version (SDQ, Goodman (2001)) | |
| | The SDQ parent questionnaire assess overall behaviour problems, emotional symptoms, hyperactivity and inattention, peer relationship problems, and prosocial behaviour. It rates participant's behaviour over the previous 6 months. The SDQ is scored on a Likert scale and includes 25 items, providing a Total Difficulties score. | SDQ total |
| | Self-reported questionnaires | |
| | KIDSCREEN-27 (Robitail et al., 2007) | |
| | The KIDSCREEN-27 is a self-reported questionnaire providing an index of health-related quality of life in | KIDSCREEN total |

| | | | |
|---|---|---|---|
| | | children and adolescents. This instrument scored on a Likert scale and includes 27 items, providing a total score. | |
| | | Social Goal Scale (SGS, Patrick, Hicks, and Ryan (1997)) | |
| | | The SGS is a self-reported questionnaire providing an index of social responsiveness and of goals setting which ultimately gets you involve with some social work. This instrument scored on a Likert scale and includes 11 items providing one score. | Social goal |
| | | Self-Compassion Scale – Short form (SCS, Raes, Pommier, Neff, and Van Gucht (2011)) | |
| | | The SCS is a self-reported questionnaire comprising 12 items, which produces a total global score that can also be classified into two subscores: negative behaviours toward the self (6 items) or positive behaviours towards the self (6 items). | Self-compassion |
| | Neuropsychological tests | | |
| | | Affect Recognition (NEPSY-II, Korkman, Kirk, and Kemp (2007) | |
| | | The affect recognition subtest assesses the ability to recognise facial emotional expressions (happy, sad, anger, fear, disgust, and neutral) from photographs of children's faces in several matching tasks. In the first task, the participant selected one of the four faces that depicted the same emotion as a child's face at the top of the page. In a second task, the participant selected two photographs of faces that displayed the same affect from a selection of four photographs. Finally, the participant examined a photograph of a child's face for 5 seconds, and then from memory, selected two photographs that matched the same emotion as the face previously shown. Standardised scores were used. | Affect recognition |
| | | Theory of Mind (NEPSY-II, Korkman et al. (2007)) | |
| | | The theory of mind subtest measures understanding of mental functions and other people's perspectives. In the first task, questions are asked to the participant about different verbal scenarios measuring understanding of beliefs, intentions, others' thoughts, ideas and comprehension of figurative language. In the second task, participants have to match facial emotional expressions, from photographs of children's faces, to a scenario. The total raw score was age-adjusted for each participant using the procedure described in the main statistical analyses section. | Theory of mind |

**Supplementary Table S2.** Randomised controlled trial timing

| | **RCT, n=56** | | |
|---|---|---|---|
| | **Intervention group (IG), n=29** | **Waiting group (WG), n=27** | **Group comparison (IG vs WG)** |
| **Difference between Time 1 and Time 2, mean (SD) in days** | 95.27 (19.9) | 90 (30.9) | t(34.73)=0.688, p=0.496 |
| **Difference between Time 2 and Time 3, mean (SD) in days** | 81.23 (21.1) | 84.64 (13.4) | t(46.98)= -0.654, p=0.502 |

Note: Independent-sample t-test was used to compare the randomised groups.

**Supplementary Table S3**. Linear model results of the intervention (IG) and wait (WG) for the delta Δ1 and Δ2 for the Intent-to-Treat Sample. Delta mean and standard deviation (SD) are also showed.

| Outcomes measures | Interaction effect time (Δ1 and Δ2) by group (IG and WG) | Planned contrast: MBI VS treatment as usual | Planned contrast: long-term VS treatment as usual | Planned contrasts Mean Δ (SD) | | |
|---|---|---|---|---|---|---|
| | | | | Treatment as usual | MBI | Long-term |
| **Executive competences** | | | | | | |
| BRIEF CEG | **F(3,80)=4.441, p=0.006, d=0.5, q=0.026** | **t(80)=-3.195,p=0.002** | t(80)= -0.589, p=0.558 | 5.16 (17.68) | -10.79 (19.43) | 1.87 (14.89) |
| BRIEF MI | **F(3,80)=6.72, p<0.001, d=0.615, q=0.006** | **t(80)= -3.985, p<0.001** | t(80)=-0.852, p=0.397 | 4.58 (12.39) | -8.31 (11.99) | 1.48 (10.49) |
| BRIEF BRI | F(3,80)=1.47, p=0.229, d=0.288, q=0.368 | t(80)= -1.327,p=0.188 | t(80)=- 0.075, p=0.941 | 0.58 (6.93) | -2.48 (9.17) | 0.39 (6.96) |
| Letter-Number sequencing | F(3,92)=0.155, p=0.962, d=0.093, q=0.962 | t(92)=0.332, p=0.74 | t(92)=0.674, p=0.502 | 0.41 (2.36) | 0.58 (1.76) | 0.81 (2.19) |
| Tempo test | F(3,91)=0.667, p=0.574, d=0.194, q=0.662 | t(91)=0.511, p=0.611 | t(91)= -0.729, p=0.468 | 4.55 (8.52) | 5.83 (8.77) | 2.42 (12.87) |
| Flanker processing speed | **F(3,90)=4.83, p=0.004, d=0.521, q=0.022** | **t(90)=-3.789, p<0.001** | **t(90)=-2.620, p=0.01** | 116.02 (256) | -140.8 (290.02) | -82.44 (180.99) |
| Flanker inhibition | F(3,90)=0.57, p=0.636, d=0.179, q=0.707 | t(90)=-0.335, p=0.739 | t(90)= 0.713, p=0.478 | -12.77 (97.33) | -26.61 (171.34) | 18.69 (141.77) |
| Reality filtering, TCC | F(3,86)=0.691, p=0.560, d=0.15, q=0.368 | t(86)=-0.795, p=0.422 | t(86)= -1.394, p=0.167 | 0.07 (0.22) | 0.04 (0.14) | 0.01 (0.1) |
| **Socio-emotional competences** | | | | | | |
| SDQ total | **F(3,88)=3.518, p= 0.018, d=0.445, q=0.048** | **t(88)=-2.423, p=0.017** | t(88)=-0.178, p=0.859 | 0.59 (3.86) | -2.02 (4.21) | 0.38 (4.18) |
| KIDSCREEN total | F(3,84)=0.401, p=0.752, d=0.15, q=0.806 | t(84)=-0.045 p=0.964 | t(84)=0.854, p=0.396 | 0.52 (13.37) | 0.39 (10.21) | 3.48 (11.64) |
| Social goal | **F(3,82)=3.812, p=0.013, d=0.463, q=0.044** | t(82)=-0.353, p=0.725 | t(82)=0.890, p=0.376 | 0.08 (0.31) | 0.04 (0.52) | 0.19 (0.41) |
| Self-compassion | F(3,82)=2.693, p=0.051, d=0.389, q=0.110 | t(82)=2.181, p=0.032 | t(82)=2.763, p=0.007 | -0.18 (0.43) | 0.15 (0.55) | 0.29 (0.66) |
| Affect recognition | F(3,92)=0.951, p=0.419, d=0.231, q=0.571 | t(92)=-0.248, p=0.804 | t(92)=-1.157, p=0.25 | 1.32 (3.31) | 1.15 (3.49) | 0.19 (3.14) |
| Theory of mind | F(3,89)=1.061, p=0.37, d=0.244, q=0.529 | t(89)=0.927, p=0.356 | t(89)=-0.546, p=0.586 | 0.52 (1.91) | 1.02 (2.08) | 0.2 (1.89) |

Abbreviations: intervention group (IG), waiting group (WG); delta scores: difference between Time 1 and Time 2 (Δ1), and between Time 2 and Time 3 (Δ2). Note: d shows Cohen's d effect sizes. All p-values of linear models that survived false discovery rate (FDR) correction according to Benjamini and Hochberg (1995) are indicated in bold (q < 0.05). Negative Δ indicate a reduction of the scores between the 2 time points whereas positive Δ indicate an increase of the scores between the 2 time points.

**Supplementary Table S4.** Planned contrast MBI VS long-term of the significant planned contrast: MBI VS treatment as usual

| Outcomes measures | Significant planned contrast: MBI VS treatment as usual (reported in supplementary table 3) | Planned contrast: MBI VS long-term |
|---|---|---|
| BRIEF CEG | **t(80)=-3.195,p=0.002** | **t(80)= -2.702, p=0.008** |
| BRIEF MI | **t(80)= -3.985, p<0.001** | **t(80)=-3.229, p=0.002** |

| | | |
|---|---|---|
| Flanker processing speed | **t(90)=-3.789, p<0.001** | t(90)= -0.926, p=0.3571 |
| SDQ total | **t(88)=-2.423, p=0.017** | **t(88)=-2.285, p=0.025** |

**Supplementary Table S5**. Subgrouping "prematurity": linear model results of the intervention (IG) and wait (WG) for the delta Δ1 and Δ2 for the Intent-to-Treat Sample, as well as delta mean and standard deviation (SD)

| Outcomes measures | Interaction effect: time (Δ1 and Δ2) by group (IG and WG) by subgrouping prematurity (high and moderate-risk) | Planned contrast: MBI VS treatment as usual | | Planned contrast: long-term VS treatment as usual | | Planned contrasts Mean Δ (SD) | | | Planned contrasts Mean Δ (SD) | | |
|---|---|---|---|---|---|---|---|---|---|---|---|
| | | | | | | Moderate-risk group | | | High-risk group | | |
| | | Moderate-risk group | High-risk group | Moderate-risk group | High-risk group | Treatment as usual | MBI | Long term | Treatment as usual | MBI | Long term |
| **Executive competences** | | | | | | | | | | | |
| BRIEF CEG | **F(7,76)=2.622, d= 0.384, p=0.018, q=0.048** | t(76)=-1.967, p=0.053 | **t(76)=-2.625, p=0.011** | t(76)=-0.911, p=0.365 | t(76)=-1.920, p=0.059 | 4.1 (14.22) | -9.55 (22.94) | -3.2 (11.22) | 6.33 (21.74) | -11.91 (16.06) | 5.77 (16.55) |
| BRIEF MI | **F(7,76)= 3.228, d=0.426, p=0.005,q=0.024** | **t(76)=-3.098,p=0.003** | **t(76)=-2.462,p=0.016** | t(76)=-1.568, p=0.121 | **t(76)=-2.216, p=0.03** | 6.5 (10.89) | -7.7 (14.02) | -1.8 (6.49) | 2.44 (14.22) | -8.86 (10.11) | 4 (12.42) |
| BRIEF BRI | F(7,76)=2.484, d=0.374, p=0.024,q=0.055 | t(76)=0.184, p=0.855 | t(76)=-2.318, p=0.023 | t(76)=0.290, p=0.773 | t(76)=-1.058, p=0.293 | -2.4 (4.72) | -1.85 (10.59) | -1.4 (7.56) | 3.89 (7.72) | -3.05 (7.88) | 1.77 (6.42) |
| Letter-Number sequencing | F(7,88)=1.567, d=0.297, p=0.156, q=0.275 | t(88)= 2.169, p=0.0328 | t(88)= -1.512, p=0.134 | t(88)=2.724, p=0.008 | t(88)=0.383, p=0.702 | -0.8 (1.55) | 0.82 (1.56) | 1.5 (2.71) | 1.42 (2.5) | 0.38 (1.92) | 0.21 (1.48) |
| Tempo test | **F(7,87)=2.944, d=0.407, p=0.008, q=0.031** | t(87)= 1.986, p=0.0501 | t(87)= -1.065, p=0.3 | t(87)=-1.558, p=0.123 | t(87)=-1.881, p=0.063 | 2.9 (9.87) | 10.19 (6.8) | -3.33 (15.29) | 5.92 (7.38) | 2.31 (8.69) | 7.36 (8) |
| Flanker processing speed | **F(7,86)=3.481, d=0.443, p=0.003, q=0.019** | **t(86)= -3.354, p=0.001** | **t(86)= -2.142, p=0.035** | **t(86)=-3.341, p=0.001** | t(86)=-1.246, p=0.216 | 250.57 (265.49) | -73.35 (219.92) | -108.78 (221.17) | -6.29 (181.86) | -200.16 (333.25) | -59.87 (142.88) |
| Flanker inhibition | F(7,86)=0.49, d=0.166, p=0.84, q=0.869 | t(86)= 0.409, p=0.683 | t(86)=-0.831, p=0.408 | t(86)=0.313, p=0.755 | t(86)=-1.516, p=0.133 | -27.43 (72.64) | -5.26 (109.19) | -7.02 (67.63) | 0.56 (117.42) | -45.39 (212.25) | 40.73 (183.44) |
| Reality filtering, TCC | F(7,82)= 0.433, d=0.215, p=0.879, q=0.662 | t(82)= -0.615, p=0.540 | t(82)= -0.499, p=0.619 | t(82)=-1.538, p=0.128 | t(82)=-0.297, p=0.767 | 0.06 (0.19) | 0.03 (0.14) | 0.03 (0.06) | 0.08 (0.26) | 0.05 (0.15) | -0.02 (0.12) |
| Socio-emotional competences | | | | | | | | | | | |
| SDQ total | F(7,84)= 1.837, d=0.321, p=0.091, q=0.170 | t(84)=-1.908, p=0.06 | t(84)=-1.509, p=0.1350 | t(84)=-0.966, p=0.337 | t(84)=-2.182, p=0.032 | 1.2 (2.53) | -1.9 (4.45) | -0.55 (4.06) | 0.08 (4.76) | -2.12 (4.09) | 1.15 (4.28) |
| KIDSCREEN total | F(7,80)=1.246, d=0.265, p=0.288, q=0.432 | t(80)=-1.454, p=0.15 | t(80)=1.161, p=0.249 | t(80)=0.076, p=0.94 | t(80)=0.097, p=0.923 | 6.89 (13.25) | 0.38 (9.7) | 7.27 (11.43) | -4.25 (11.79) | 0.39 (10.87) | 0 (11.17) |
| Social goal | F(7,78)=1.989, d=0.334, p=0.067, q=0.134 | t(78)= -0.482, p=0.631 | t(78)= -0.085, p=0.932 | t(78)=0.756, p=0.452 | t(78)=0.421, p=0.675 | 0.16 (0.39) | 0.09 (0.58) | 0.31 (0.32) | 0.01 (0.22) | -0.01 (0.46) | 0.09 (0.46) |
| Self-compassion | **F(7,78)=6.135, d=0.587, p<0.001, q<0.001** | t(78)= 0.540, p=0.591 | **t(78)= 3.015, p=0.004** | **t(78)=3.165, p=0.002** | **t(78)=2.743, p=0.008** | -0.02 (0.38) | 0.07 (0.64) | 0.67 (0.64) | -0.32 (0.43) | 0.22 (0.46) | -0.06 (0.48) |
| Affect recognition | F(7,88)=0.823, d=0.215, p=0.571, q=0.662 | t(88)=0.459, p=0.648 | t(88)=-0.751, p=0.454 | t(88)=-1.060, p=0.292 | t(88)=0.447, p=0.656 | 0.7 (2.36) | 1.32 (2.03) | -0.83 (1.7) | 1.83 (3.97) | 1 (4.41) | 1.07 (3.83) |
| Theory of mind | F(7,85)= 0.887, d=0.223, p=0.521, q=0.662 | t(85)=0.910, p=0.365 | t(85)=0.457, p=0.649 | t(85)=-0.682, p=0.497 | t(85)=0.282, p=0.779 | 0.6 (2.55) | 1.33 (2.11) | 0 (1.48) | 0.45 (1.21) | 0.77 (2.07) | 0.36 (2.21) |

Abbreviations: intervention group (IG), waiting group (WG); delta scores: difference between Time 1 and Time 2 (Δ1), and between Time 2 and Time 3 (Δ2). Note: d shows Cohen's d effect sizes. All p-values of linear models that survived false discovery rate (FDR) correction according to Benjamini and Hochberg (1995) are indicated in bold (q < 0.05). Negative Δ indicate a reduction of the scores between the 2 time points whereas positive Δ indicate an increase of the scores between the 2 time points.

**Supplementary Table S6**. Subgrouping "prematurity": Planned contrast MBI VS long-term of the significant planned contrast: MBI VS treatment as usual

| Outcomes measures | Significant planned contrast: MBI VS treatment as usual (reported in supplementary table 5) | | Planned contrast: MBI VS long-term | |
|---|---|---|---|---|
| | Moderate-risk group | High-risk group | Moderate-risk group | High-risk group |
| BRIEF CEG | t(76)=-1.967, p=0.053 | **t(76)=-2.625, p=0.011** | | **t(76)= -2.879, p=0.005** |
| BRIEF MI | **t(76)=-3.098,p=0.003** | **t(76)=-2.462,p=0.016** | t(76)= -1.287 0.2019 | **t(76)= -3.159, p=0.002** |
| Flanker processing speed | **t(86)= -3.354, p=0.001** | **t(86)= -2.142, p=0.035** | t(86)= 0.416 0.6782 | t(86)= -1.682, p=0.096 |
| Self-compassion | t(78)= 0.540, p=0.591 | **t(78)= 3.015, p=0.004** | | t(78)= 1.602, p=0.113 |
